# EASILY SCALABLE, RAPIDLY DEPLOYABLE MECHANICAL VENTILATOR FOR PANDEMIC HEALTH CRISES IN RESOURCE-LIMITED AREAS

**DOI:** 10.64898/2026.04.08.26350386

**Authors:** Ramon Farré, Raffaella Salama, Miguel A. Rodríguez-Lázaro, Kasra Kiarostami, Laia Fernández-Barat, Viviane De Cassia Oliveira, Antoni Torres, Núria Farré, Anh Tuan Dinh-Xuan, David Gozal, Jorge Otero

**Affiliations:** Unitat de Biofisica i Bioenginyeria, Facultat de Medicina i Ciències de la Salut, Universitat de Barcelona, Spain; CIBER de Enfermedades Respiratorias, Spain; Institut Investigacions Biomèdiques August Pi Sunyer, Barcelona, Spain; Portiuncula University Hospital and East Galway and Roscommon Integrated Care Hub, Ballinasloe, Ireland; School of Medicine, University of Galway, Galway, Ireland; Department of Respiratory Medicine and Physiology, Hôpital Cochin, Paris, France; Departments of Pediatrics and Biomedical Sciences and Office of the Dean, Joan C. Edwards School of Medicine, Marshall University, Huntington, WV, USA

**Keywords:** Mechanical ventilation, respiratory pandemics, emergency, low-resource, open-source, last-resort therapy

## Abstract

**Background:** The COVID-19 pandemic exposed critical shortages of mechanical ventilators, particularly in low-resource settings. Disruptions in global supply chains and dependence on specialized components highlighted the need for scalable, locally manufacturing alternatives for emergency respiratory support.

**Aim:** To describe and evaluate a simplified, supply-chain-independent mechanical ventilator assembled from widely available automotive and simple hardware components, and intended as a last-resort solution.

**Methods:** The ventilator is based on a reciprocating air pump driven by an automotive windshield wiper motor coupled to parallel shaft bellows and readily assembled passive membrane valves, only requiring materials available from standard hardware retailers, minimal tools, and basic manual skills. Ventilator performance was assessed through bench testing using a patient model simulating severe lung disease in an adult (R=20 cmH₂O·s/L, C=15 mL/cmH₂O) and pediatric (R=50 cmH₂O·s/L, C=10 mL/cmH₂O) patients. Realistic proof of concept was performed in four mechanically ventilated 50-kg pigs.

**Results:** The device delivered tidal volumes up to 600 mL and respiratory rates up to 45 breaths/min with PEEP up to 10 cmH₂O, covering pediatric and adult ventilation ranges. In vivo testing showed that the ventilator maintained arterial blood gases within the targeted range. Technical details for ventilator construction are provided in an open-source video tutorial.

**Discussion:** This low-cost ventilator demonstrated adequate performance under demanding conditions. Although not a substitute for commercial intensive care ventilators, its simplicity, autonomy, and independence from fragile supply chains provide a potentially life-saving option in resource-constrained emergency scenarios.

## INTRODUCTION

The COVID-19 pandemic exposed the world to an unprecedented health crisis, resulting in a high mortality (1,2) primarily driven by respiratory failure (3). The scale and severity of the disease fostered an extraordinary demand for mechanical ventilators that exceeded the capacity of even the most advanced healthcare systems (4,5). This situation necessitated urgent modifications to medical device regulations and exceptional procedures to facilitate the provision of mechanical ventilation and enable survival among all severely affected patients (6–10). This recent health crisis has made it clearer than ever that the world remains at constant risk of infectious pandemics –whether due to new influenza variants, emerging coronaviruses or other (11,12)– that primarily affect the lungs and require respiratory intensive care. The relevant question is no longer whether another such pandemic will occur, but when it will emerge and spread globally, underscoring the urgent need for appropriate preparedness strategies (11).

The experience of the COVID-19 pandemic drove high-income countries to implement strengthened preparedness strategies, including stockpiling of ventilators and associated consumables, as well as the development of contingency plans for rapid manufacturing. Unfortunately, the issue of preparedness remains virtually unresolved in low- and middle-income countries, where baseline shortages in critical care resources persist even in the absence of a sudden emergency (13). During a respiratory pandemic surge, low-resource regions can scarcely rely on large-scale provision of ventilators from high-income countries. Even in the absence of financial constraints, health systems in developed nations are themselves likely to be overwhelmed, limiting their capacity to supply external support. Furthermore, the feasibility of locally manufacturing ventilators, based on conventional technology or simplified innovations proposed during the COVID-19 pandemic (14,15), remains limited in the most deprived settings. Indeed, such initiatives typically require a baseline level of technological infrastructure and specialized expertise that is often unavailable in resource-constrained regions. Moreover, global health crises profoundly disrupt international supply chains for critical technical components, as evidenced during the COVID-19 pandemic (16), thereby restricting access to essential parts required for ventilator assembly.

Under these circumstances, a more realistic strategy for expanding urgent access to mechanical ventilation in low-resource settings during a pandemic would involve the development of a scalable and rapidly deployable mechanical ventilator platform. Such a system should be designed to allow local device assembly by available personnel and constructed from materials that are readily accessible across diverse settings. Importantly, this approach should avoid dependence on fragile global supply chains for specialized technological and medical components, thereby enhancing resilience during periods of global disruption. Accordingly, we here describe a solution that enables the simple construction of a performant ventilator using widely available components. By returning to core physiological and engineering concepts, the proposed ventilator minimizes technological complexity while preserving the essential functions required to support severely affected patients. Following an open-source publication approach (17,18), the device’s construction process is described in detail to facilitate reproducibility and local implementation. The ventilator performance was rigorously evaluated through bench testing under strenuous conditions, followed by preclinical assessment in a porcine model. Remarkably, the ventilator presented herein is not intended to replace commercially available intensive care equipment. Rather, it is conceived as a last-resort alternative designed to provide potentially lifesaving ventilatory support in contexts where no other alternatives are available. In settings where access to mechanical ventilation is otherwise unattainable, such a solution may substantially reduce preventable mortality during respiratory pandemics.

## METHODS

### Ventilator description

The proposed ventilator is based on a classical mechanical configuration using a reciprocating air pump and inspiratory and expiratory valves (Figure 1). The originality of the device presented herein lies in that it requires only common and particularly cheap automotive parts and other simple materials available from standard hardware retailers, minimal tools, and basic manual skills. The ventilator is briefly described below, and full details and construction instructions are provided by a video tutorial in the *Supplementary Materials*.

**Figure 1.**
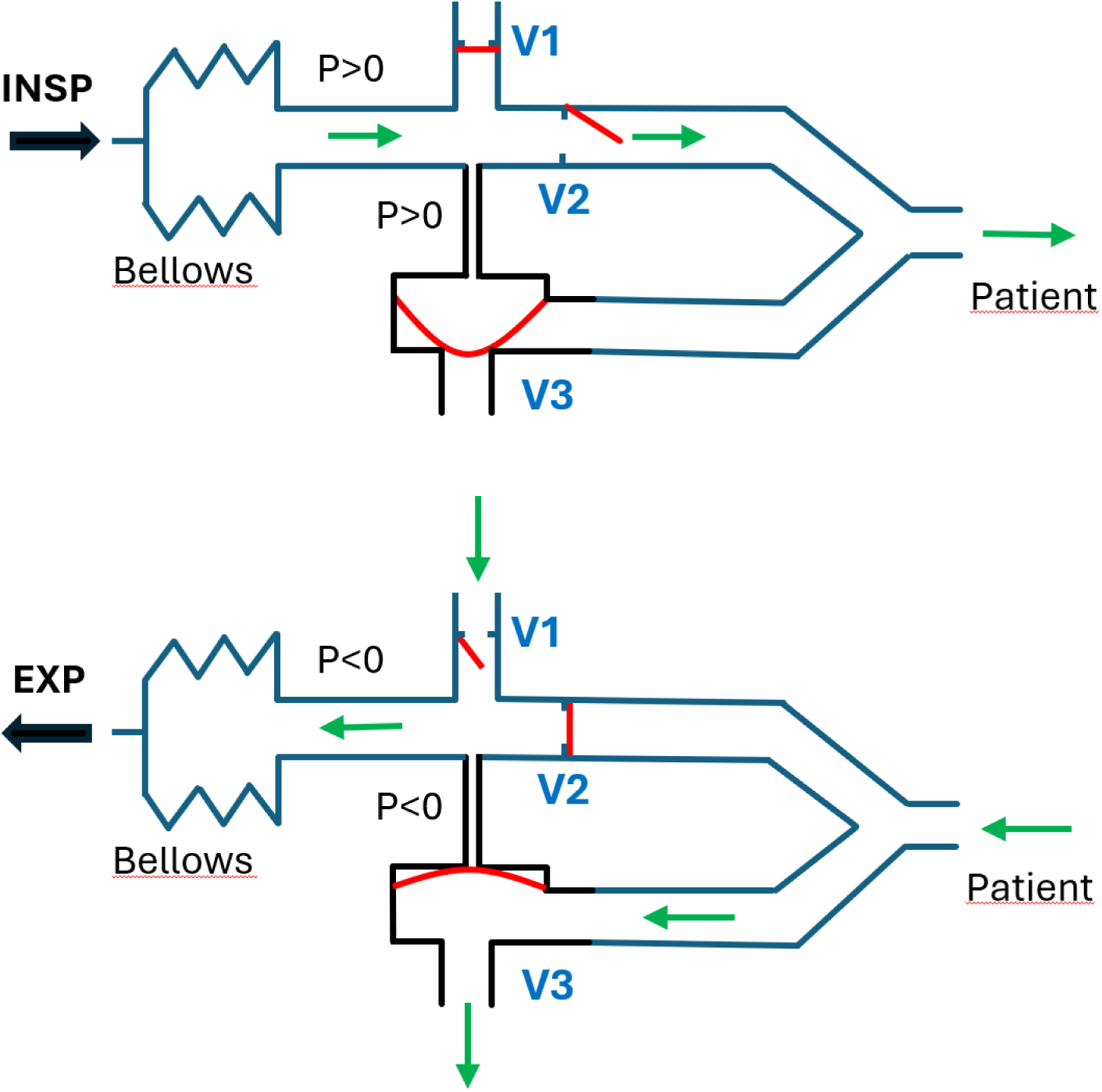
Ventilator diagram showing the open or closed status of valves V1, V2 and V3 during inspiration (INSP) and expiration (EXP). Flaps in valves V1 and V2 and membrane in valve V3 are shown in red. Black arrows indicate bellows movement. Green arrows indicate air flow.

The cycling air pump (Figure 1) is actuated by a standard car windshield wiper motor (Figure 2) which directly provides reciprocating linear displacement and typically operates at 40–60 cycles per minute when powered by a 12-V car battery. A simple DC motor speed regulator, commonly available as an inexpensive device for controlling automotive fan speed, is used to adjust the cycling frequency. This allows the respiratory rate to be set within the range required for mechanical ventilation in both pediatric and adult patients (12–45 breaths/min).

**Figure 2.**
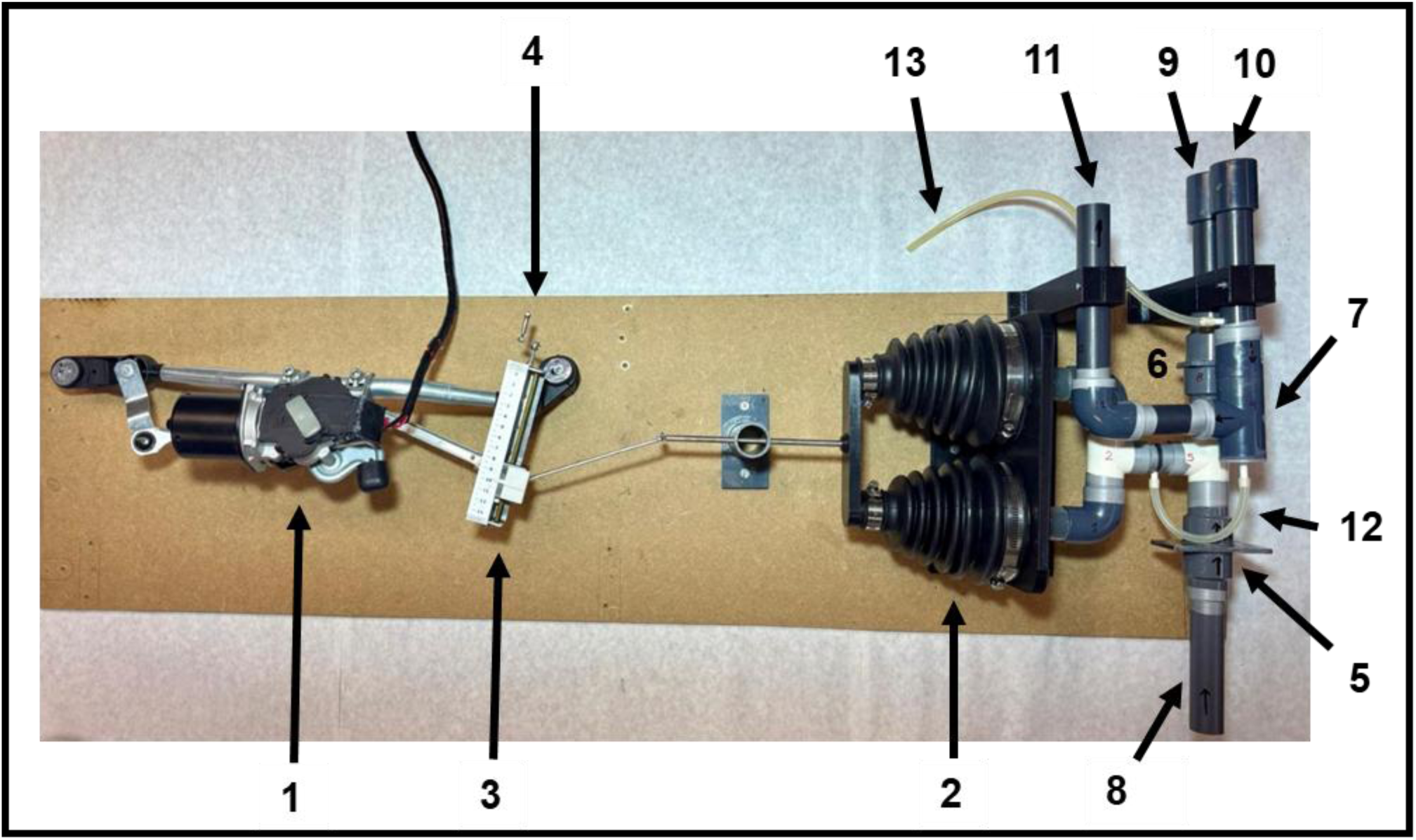
Connection of the ventilator basic components: windshield wiper motor (1), drive shaft bellows (2), variable-length adjustment mechanism (3) including a crank for adjusting bellows displacement (4), flap valves in the inspiratory lines (5, 6), expiratory valve (7), room/O2 air inlet (8), connector for inspiratory (9) and expiratory (10) tubing to the patient’s airway, expiratory outlet (11) for connection to a PEEP bottle, tube to drive the expiratory valve by pump pressure (12), tube for sensing airway pressure with a U-tube manometer (13). Detailed construction instructions are provided by a video tutorial in Supplementary materials.

The reciprocating motor is used to drive shaft bellows (Figure 2) which are widely used in most car models to protect boots that surround drive shaft joints. To achieve the tidal volume (V_T_) required for adult ventilation, two such bellows, sealed at both ends with caps, are mounted in parallel and actuate simultaneously using a shared mechanical linkage. A rigid connector links both bellows to the actuation mechanism (Figure 2), ensuring balanced force distribution during compression and synchronized movement. The windshield wiper motor is mechanically connected to the moving end of the bellows via a rod that produces linear back-and-forth to drive the air pump (Figure 2). The connecting rod incorporates a variable-length adjustment mechanism that allows manual modification of the bellows displacement amplitude (and thus V_T_) by means of a crank, without interrupting ventilation (Figure 2). This adjustment system is constructed using commonly available hardware materials, including a threaded rod and nuts.

The ventilator valves are passively operated by the reciprocating pump and therefore do not require active control elements (Figure 1): V1 and V2 are flap (i.e., differential pressure-driven) valves, and V3 is based on a flexible membrane passively controlled by the bellows compartment pressure (Figure 1). These valves are manufactured using simple and widely available materials. V1 and V2 membranes are cut from plastic folder sheets, while the flexible membrane for V3 is cut from latex or nitrile clinical gloves. Both valve types are assembled using PVC pipes and fittings (Figure 2). The air inlet valve (V1) allows direct intake of room or oxygen-enriched air (Figure 2). Positive end-expiratory pressure (PEEP) is set using the classical method of immersing the expiratory outlet tube into a water column at a height corresponding to the target PEEP (Figure 3).

**Figure 3.**
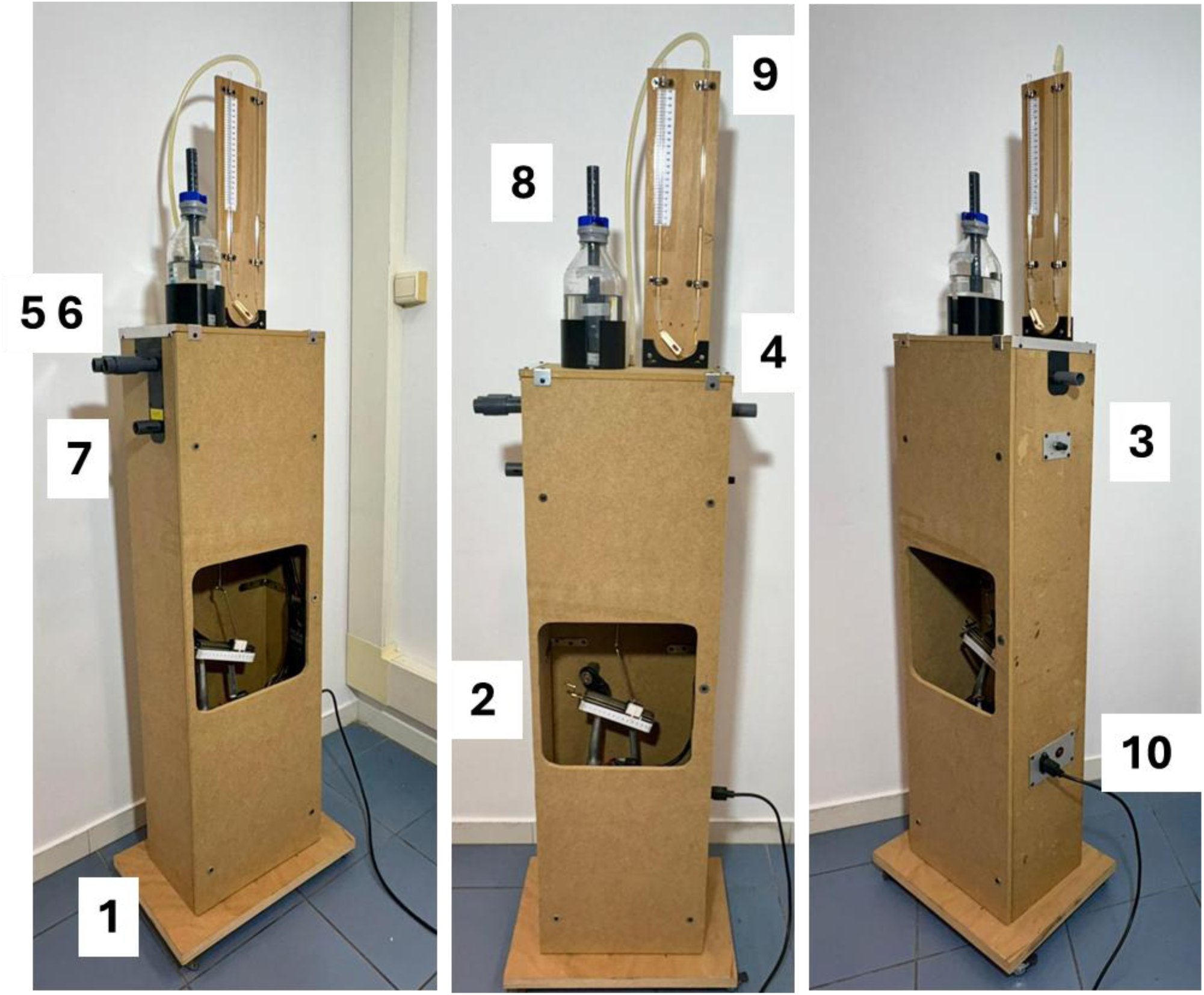
Assembled ventilator on a stand-alone platform with wheels (1). The platform in Figure 2 is placed vertically and enclosed in a wooden box: window for accessing the crank for modifying tidal volume (2), button of the frequency regulator to set breathing rate (3), room/O2 air inlet (4), connector for inspiratory (5) and expiratory (6) tubing to the patient’s airway, expiratory outlet (7) for connection to PEEP bottle (8), U-tube manometer for assessing airway pressure (9), electrical power connection (10) (12-V car battery or conventional 110/220 V AC-line).

A water-filled transparent U-tube (5-mm ID, 45-cm arm length) connected to the inspiratory outlet allows continuous monitoring of airway pressure, particularly peak inspiratory pressure, which is a key parameter for quantifying the magnitude of mechanical ventilation delivered to the patient (Figure 3). An adjustable resistance (constructed from the roller clamp of a conventional intravenous infusion set) is placed between the two arms of the U-tube to optimize the dynamic response of the manometer at ventilation frequencies (Figure 3). Importantly, the U-tube manometer also functions as a safety valve, preventing dangerous lung overpressure in the event of expiratory valve failure resulting in persistent closure during expiration.

### Technical performance: bench test

Ventilator bench testing was performed using a physical model that mimics the mechanical properties of the respiratory system (SmartLung Adult, IMT Analytics, Switzerland), consisting of adjustable resistance (R) and compliance (C) elements (13). These selectable parameters allowed simulation of a wide range of respiratory mechanical conditions representative of different patient populations. Flow and pressure were measured at the entrance of the respiratory model using a pneumotachograph and a pressure transducer, respectively. The electrical current supplied to the ventilator, and therefore its power consumption, was measured. After more than 400 h of continuous ventilation of a patient model, the prototype was tested under conditions simulating healthy lungs and, more importantly, under demanding conditions corresponding to severely diseased adult (R=20 cmH₂O·s/L, C=15 mL/cmH₂O) and pediatric (R=50 cmH₂O·s/L, C=10 mL/cmH₂O) patients. The respiratory rate was set up to 24 breaths/min and up to 45 breaths/min for adult and pediatric simulations, respectively. PEEP levels of up to 10 cmH₂O were applied. The capacity of the ventilator for gas exchange was tested on the bench by simulating physiological CO_2_ production and observing how the ventilator was able to keep normocapnia. To this end, a known continuous flow of CO_2_ (ranging 45–350 mL/min) was injected into the compliant component of the patient model and end-tidal CO₂ (EtCO₂) was assessed by capnometry (Capnostream 20p, Oridion) at the airway opening (Y-piece of ventilator tubing).

### *In vivo* performance: porcine ventilation

After bench testing, proof of concept for ventilator performance was evaluated in four adult pigs (48–52 kg) following a protocol approved by the competent ethical board. As detailed in the *Supplementary Materials*, the animals were initially instrumented and ventilated using a conventional setting. Once stable physiological conditions were achieved, the commercial ventilator was replaced by the prototype device, with its admission valve V1 connected to an O_2_ source through a Venturi valve (from a mask setting) to provide FiO_2_=0.4. From initial settings V_T_=8 ml/kg, PEEP=5 cmH_2_O and 20 breath/min, the ventilator was adjusted (mainly breathing rate) as required to maintain arterial PCO_2_ within normal values (35-45 mmHg). After 40 minutes of stable ventilation with the prototype ventilator, physiological data were recorded, and arterial blood samples were collected for gas analysis. The animals were finally euthanized with i.v. infusion of an overdose of sedatives and KCl (1 mEq/kg).

## RESULTS

### Bench Performance Under Severe Mechanical Conditions

The ventilator was performed satisfactorily when used in patient models characterized by markedly increased resistance and reduced compliance. When tested on an adult obstructive–restrictive patient model (R=20 cmH₂O·s/L, C=15 mL/cmH₂O), the ventilator delivered V_T_ of up to 600 mL, sufficient to provide protective ventilation (6 ml/kg) for an 85 kg patient. Figure 4 (left) shows representative pressure and flow signals recorded while ventilating this adult model (24 breaths/min, V_T_=400 mL) and a high-resistance, low-compliance pediatric model (R=50 cmH₂O·s/L, C=10 mL/cmH₂O) at a rate of 45 breaths/min with V_T_=90 mL (which would correspond to patients with body weight down to <10 kg). The inspiratory flow exhibited a sinusoidal-like pattern quite similar to the physiological inspiratory flow and consistent with the mechanical design of the ventilator air pump. Expiration showed the typical passive flow decay observed in patients. Figure 4 (right) shows the corresponding signals recorded when the same adult and pediatric models were ventilated with PEEP=10 cmH_2_O. In this case, the expiratory portions of both pressure and flow signals displayed slight oscillations caused by bubble formation as PEEP was generated. Electrical power consumption of the ventilator was 7.3 W (PEEP=0) and 12.0 W (PEEP=10 cmH₂O) for the adult model, and 18.5 W (PEEP=0) and 20.1 W (PEEP=10 cmH₂O) for the pediatric model. By adjusting the ventilation amplitude and frequency within physiological ranges (16-26 breaths/min and 120-500 mL, respectively) the ventilator maintained a normal value of EtCO_2_=35 mmHg for CO_2_ productions up to 350 mL/min, thus covering the metabolic activity of pediatric and adult patients.

**Figure 4.**
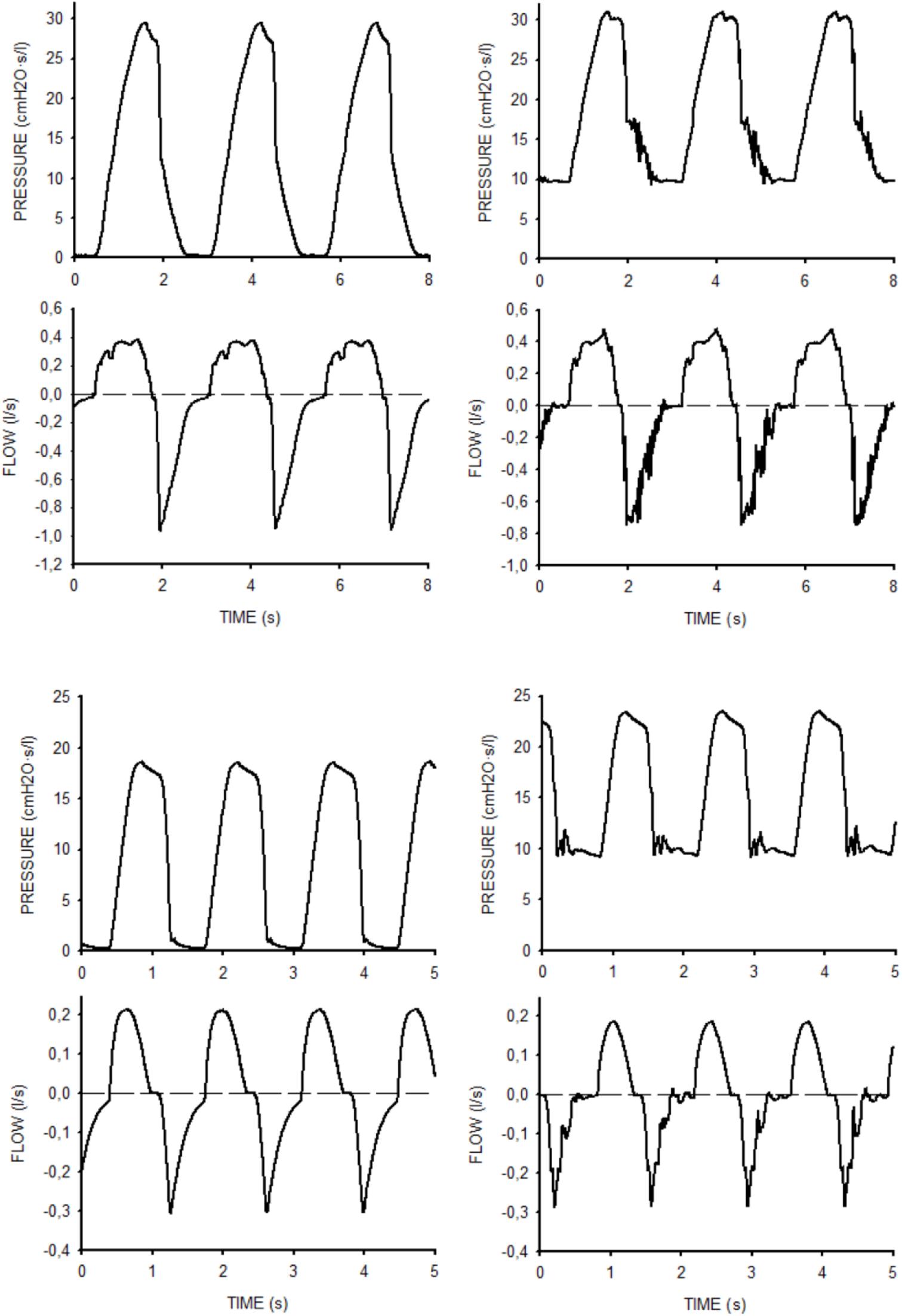
Examples of the ventilator performance in the bench test on an adult obstructive-restrictive patient model (top) and on a child patient model (bottom), without (left) and with (right) PEEP=10 cmH_2_O. Pressure and flow (positive during inspiration) were measured at the airway opening of the patient model.

As illustrated by Figure 5, the simple U-tube manometer served as an effective safety valve. After the expiratory outlet of the ventilator was intentionally occluded, when airway pressure reached the level corresponding to the height of the U-tube column, its water was expelled, thereby limiting inspiratory pressure. This mechanism prevented progressive lung overdistension that would otherwise have resulted from continued ventilator cycling while the expiratory valve was abnormally occluded.

**Figure 5.**
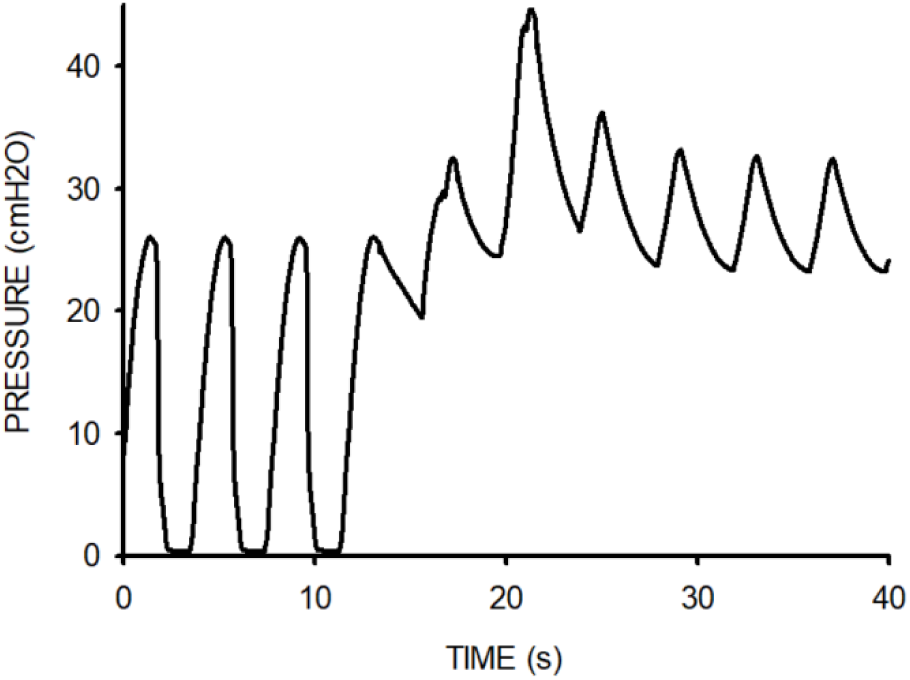
Airway pressure recorded during mechanical ventilation when, in the bench test, the expiratory outlet of the ventilator was kept intentionally closed (starting at time ≈12 s). Airway pressure increased to only 44 cmH_2_O transiently, avoiding lung over-ventilation.

### *In Vivo* Performance: Porcine Ventilation

As expected from the bench testing results, the ventilator was able to easily and satisfactorily ventilate the 50-kg pigs. With ventilator settings (m±SD) V_T_=391±7 ml, 19.3±2.1 breath/min and PEEP=5.5±0.5 cmH_2_O, physiological parameters were arterial PCO_2_=38.3±2.1 mmHg, arterial PO_2_=188.3±25.0 mmHg (FiO_2_=0.4), heart rate = 70.7±6.8 bpm, and arterial pressure (max / min)= 137.7±6.8 / 81.3±3.1 mmHg. Figure 6 shows representative recordings of airway pressure and flow during ventilator setting in one of the pigs (51 kg; V_T_=395 ml). PCO_2_ was 44 and 35 mmHg for 18 and 23 breaths/min, respectively. Thus, ventilation frequency for this animal was set to 20 breaths/min (PCO_2_ = 40 mmHg).

**Figure 6.**
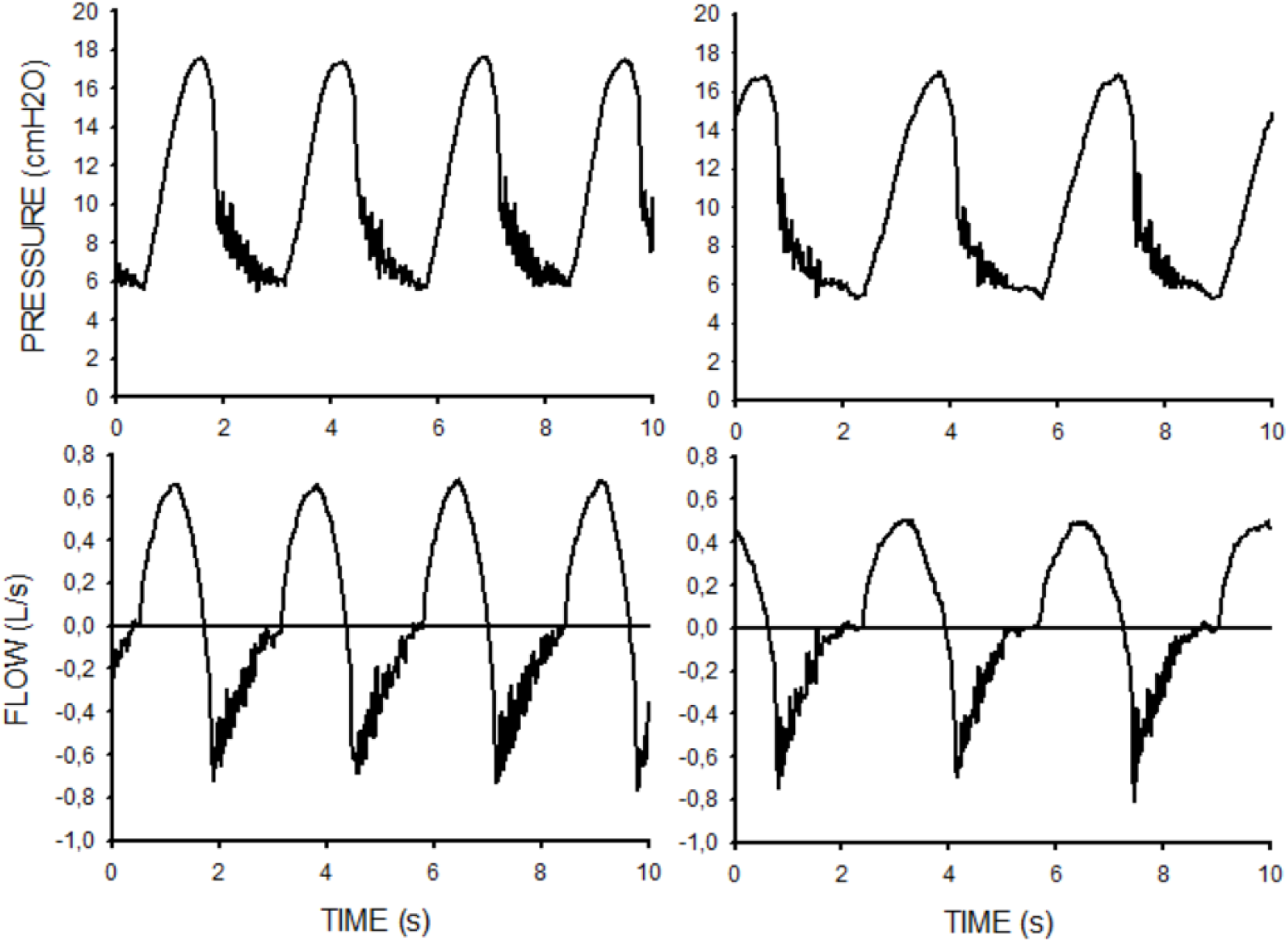
Airway pressure and flow (positive during inspiration) recorded when the prototype ventilator was used in a representative pig, with frequencies of 23 breaths/min (left) and 18 breaths/min (right).

## DISCUSSION

This study was not intended to develop a state-of-the-art ventilator, but rather to design a performant life-saving device easily assembled from components that are widely available across diverse settings, particularly in low-resource environments. To minimize structural and functional complexity, the device is based on early ventilator designs. Accordingly, the device obviates the need for a compressed air source, specialized high-pressure blowers, electronic sensors, commercialized valves and complex control systems. Its technical performance is adequate to deliver ventilatory support across a broad range of respiratory frequencies (up to 45 breaths·min⁻¹) and tidal volumes (up to 600 mL), thereby covering patients from pediatric to adult body weight categories. In addition to comprehensive bench testing, poor-of-concept of the device was carried out in a porcine model, demonstrating satisfactory functional performance *in vivo*.

The ventilator is constructed around a mechanically driven airway pump coupled with passive membrane-based valves. As the motion-generating element, a standard automotive windshield wiper motor was selected due to its widespread availability and integrated mechanical features. Indeed, this component already incorporates a rotary motor, reduction gear assembly, and mechanical linkage, enabling direct generation of reciprocating motion suitable for bellows actuation in terms of both displacement amplitude and cycling frequency. Ventilatory rate adjustment is achieved through a simple passive regulator, while tidal volume is modulated via a selectable mechanical linkage that modifies stroke amplitude. Notably, windshield wiper motors are engineered for sustained operation under demanding conditions, are powered by a conventional 12-V automotive battery and exhibit low energy consumption. Under high-demand ventilation settings, electrical power measurements in the bench test indicated that the charge stored in a standard 12-V battery car (e.g., 80 A·h) would sustain continuous ventilation for approximately 5.5 and 3.4 days in an adult patient without and with PEEP, respectively. Corresponding autonomy for pediatric ventilation was estimated at 2.2 and 2.0 days, respectively.

Shaft bellows were selected as the air-displacement element because they constitute a widely available automotive component suitable for ventilator construction. These bellows are typically manufactured from impermeable, flexible, and durable rubber or synthetic polymers engineered to withstand repetitive mechanical deformation under sustained operating conditions. The prototype incorporated two bellows arranged in parallel to achieve the desired volumetric output. However, depending on the specific bellows employed or the target maximum effective tidal volume, the number of parallel units can be increased or reduced to adjust volumetric capacity. Given that for the pressures involved in mechanical ventilation (<40 cmH_2_O) the bellows are virtually incompressible, and thus behave as a piston in a cylinder, the crank-displacement scale is useful for adjusting V_T_.

The ventilator valves were designed for ease of manufacture, drawing on early ventilatory valve concepts, many of which are currently employed in routine clinical devices such as manual resuscitation bags (19). Flap valves were selected for V1 and V2, as their functional requirement is straightforward: during inspiration –when positive pressure is generated within the bellows– they close and open, respectively; during expiration –when negative pressure develops within the bellows– they open and close, respectively. In contrast, valve V3 requires a different operational principle. This valve must remain closed during inspiration and open during expiration, independently of the pressure gradient between the patient airway and the ventilator outlet during either phase of the respiratory cycle. To fulfill this requirement, V3 was constructed based on early ventilation valve designs (20,21), consisting of a flexible membrane passively actuated by pressure changes within the bellows chamber (Figure 1). During inspiration, positive pressure within the bellows compartment displaces the V3 membrane to occlude the expiratory limb. Conversely, during expiration, negative pressure within the bellows compartment repositions the membrane, thereby opening the expiratory pathway.

One notable simplification feature of the ventilator is the absence of electronic sensors for monitoring respiratory variables. Instead, the system incorporates a simple U-tube manometer to continuously assess airway pressure (e.g., peak and end-inspiratory values), which is relevant for ventilation monitoring. Importantly, the manometer also enables continuous verification of proper valve functioning, thereby contributing to patient safety. Indeed, if during the inspiratory phase the pressure displayed on the manometer remains close to zero, this finding may indicate one of the following abnormalities: (i) valve V1 remaining open, allowing the air displaced by the bellows to leak into the ambient environment; (ii) valve V2 remaining closed, preventing airflow delivery to the patient; or (iii) valve V3 remaining open, resulting in inspiratory air preferentially escaping to the environment through the expiratory limb. Airway pressure would also remain negligible if V1 is erroneously closed during expiration. In this scenario, ambient air cannot refill the bellows, thereby precluding delivery of the subsequent inspiratory cycle. Interestingly, the manometer may additionally detect malfunctions associated with potential rebreathing. For example, abnormal persistence of V2 in the open position during expiration would allow expired gas to enter the bellows. This malfunction would be readily identified by the appearance of negative pressure during the expiratory phase. Finally, as previously illustrated (Figure 5), the manometer also functions as a safety valve in the event of inappropriate closure of V3 during expiration.

During the COVID-19 pandemic, numerous simplified versions of emergency ventilators were proposed although only a limited proportion underwent *in vivo* validation (15). While several of these designs represent thoughtful device simplifications with the potential to support safe mechanical ventilation, the majority rely on compressed air source or electronic and medical-grade components sourced from conventional commercial supply chains (e.g., high-pressure blowers, transducers, resuscitation bags, inspiratory/expiratory and PEEP valves). Although such dependence is not problematic under routine circumstances, it constitutes a critical vulnerability during global health emergencies, when supply chains are disrupted (16). The proposed ventilator addresses this limitation through a fundamentally different design strategy. Its fabrication does not depend on specialized technological or medical supply chains. Instead, it utilizes low-cost, locally available mechanical components derived from the automotive sector. Given the near-ubiquitous global presence of motor vehicles and their associated components, this approach leverages an existing, decentralized industrial infrastructure. Furthermore, the ventilator’s open modular architecture enables rapid and straightforward replacement of individual components using spare parts. As a result, this low-cost device offers a potentially scalable and supply-resilient alternative for emergency ventilatory support in resource-constrained or crisis settings.

Historical experience demonstrates that, in large-scale health emergencies, life-sustaining therapies and medical devices are often deployed before comprehensive validation or formal regulatory approval, signaling operational necessity of improvisation when conventional well-established treatments are insufficient or unavailable (22–25). Referring specifically to medical devices in respiratory emergencies, during mid-20th century poliomyelitis epidemics, negative-pressure ventilators and manually operated bag ventilation were implemented at scale despite limited standardization, successfully prioritizing functional respiratory support over technological refinements (26). The COVID-19 pandemic is a more recent example where exceptional actions were taken regarding the usability of mechanical ventilators (5,6). These precedents demonstrate a consistent general principle in disaster medicine: when mortality risk is immediate and infrastructure is overwhelmed, the threshold for deploying simplified, investigational, or non-conventional technologies shifts toward pragmatic functionality and scalability (27–31). Within this framework, the exceptional use of the ventilator described here, even though it may be previously unregulated and built with non-medical grade components, would align with established vital response paradigms (beneficence, non-maleficence and scientific plausibility) if permission from the local authority in charge of the emergency is granted (30,31).

In conclusion, in the context of a health emergency by respiratory pandemic in a low-resource setting, the proposed supply-chain-independent ventilator, or a slight variant following the same design strategy, may provide a last-resort tool to provide respiratory support to patients who otherwise would have no access to this life-saving therapy.

## Data Availability

All data produced in the present work are contained in the manuscript

## SUPPLEMENTARY MATERIAL

The Supplementary materials contain a “Supplementary methods” file and a “Video Tutorial” with detailed instructions for constructing the ventilator.

## ACKNOWLEDGMENTS

This work was partially supported by Sociedad Española de Neumología y Cirugía Torácica (SEPAR) (grant 1381/2022). SEPAR had no involvement other than providing funding for the independently submitted research project.

## ETHICAL PERMISSION

The specific animal research protocol followed in this study was approved by the Animal Research Ethics Committee of the University of Barcelona (permission number 694/25), in compliance with the relevant European and National regulations.

## AUTHORS’ CONTRIBUTION

R. Farré conceived the project, supervised the study and wrote the manuscript. R. Salama, M.A. Rodriguez-Lazaro and J. Otero contributed to the construction and bench testing of the ventilator. K. Kiarostami, L. Fernandez-Barat and V.D.C. Oliveira participated in the animal experiments. A. Torres, N. Farré, A.T. Dinh-Xuan and D. Gozal contributed to the physiological and clinical contextualization of the study. All authors revised the manuscript and agreed to its final version.

## USE OF ARTIFICIAL INTELIGENCE

Artificial intelligence (ChatGPT, free version) was used only to check the wording of a preliminary version of the manuscript.

## VIDEO TUTORIAL

To directly see (no need to previously download): https://ubarcelona-my.sharepoint.com/:v:/g/personal/raffaellasalama_ub_edu/IQCmbDKw6cuhRaD4brloZkIJAefdIU83A2ejbftvEum_oS8?nav=eyJyZWZlcnJhbEluZm8iOnsicmVmZXJyYWxBcHAiOiJPbmVEcml2ZUZvckJ1c2luZXNzIiwicmVmZXJyYWxBcHBQbGF0Zm9ybSI6IldlYiIsInJlZmVycmFsTW9kZSI6InZpZXciLCJyZWZlcnJhbFZpZXciOiJNeUZpbGVzTGlua0NvcHkifX0&e=sOc3ug

To download from a permanente public repository: https://doi.org/10.5281/zenodo.19467677

## METHODS

### Ventilator description

#### Criterion for materials selection and assembly

The material selection strategy prioritized availability, affordability, and ease of fabrication in order to facilitate deployment in low-resource settings. To emulate the operational constraints expected during pandemic conditions, the prototype was deliberately constructed without the use of specialized materials, precision-manufactured components, or advanced machining processes. Instead, widely available off-the-shelf components were employed, including standard wooden structural elements, PVC tubing, commercial adhesives, and other parts commonly obtainable from hardware suppliers. Fabrication and assembly were performed using basic workshop tools –such as manual saws, files, and drills– thereby minimizing dependence on specialized manufacturing infrastructure. Nevertheless, the proposed design is modular and can be readily upgraded, depending on local technical capabilities and component availability, to achieve improved mechanical robustness and a more industrial-grade appearance.

#### Ventilator assembly

The ventilator architecture is primarily composed of an air pump and a valve system responsible for air flow control. The air pump is based on an automotive windshield wiper motor coupled to a bellows-type air chamber through a mechanical transmission mechanism that enables adjustment of the delivered tidal volume. The specific prototype developed in this study is illustrated in the video tutorial provided in the Supplementary Materials. Since windshield wiper motors and bellows assemblies from different vehicle models may be used, the geometric configuration and dimensions of the mechanical assembly can be easily adapted. The video tutorial also demonstrates the modular nature of the system design, which allows straightforward replacement of any individual component.

#### U-tube manometer

The U-tube manometer consists of two vertical liquid-filled columns connected at their lower ends. When a differential pressure is applied between the open ends of the two columns, the resulting displacement of the liquid column is proportional to the pressure difference. The dynamic behavior of the U-tube manometer is governed by the mechanical balance between a conservative component, represented by the inertial mass of the moving liquid column, and a dissipative component, determined by viscous flow resistance within the system (1). For a U-tube with an internal diameter of 5 mm and an effective liquid column height of 40 cm, the system behaves as a second-order underdamped oscillator with a resonance frequency of approximately 60 cycles/min, which is higher than, but relatively close to, typical mechanical ventilation frequencies. Therefore, to optimize dynamic performance for airway pressure monitoring at clinically relevant ventilation rates, the U-tube manometer should be adjusted to achieve critical damping (1). Damping can be increased by augmenting the viscous component of the system through the introduction of a flow resistance between the two branches of the U-tube (2). A practical and readily available solution consists of incorporating the roller clamp of a conventional intravenous infusion set into the connecting segment (Figure 1.A), thereby providing a simple and adjustable resistance mechanism.

**Figure 1.**
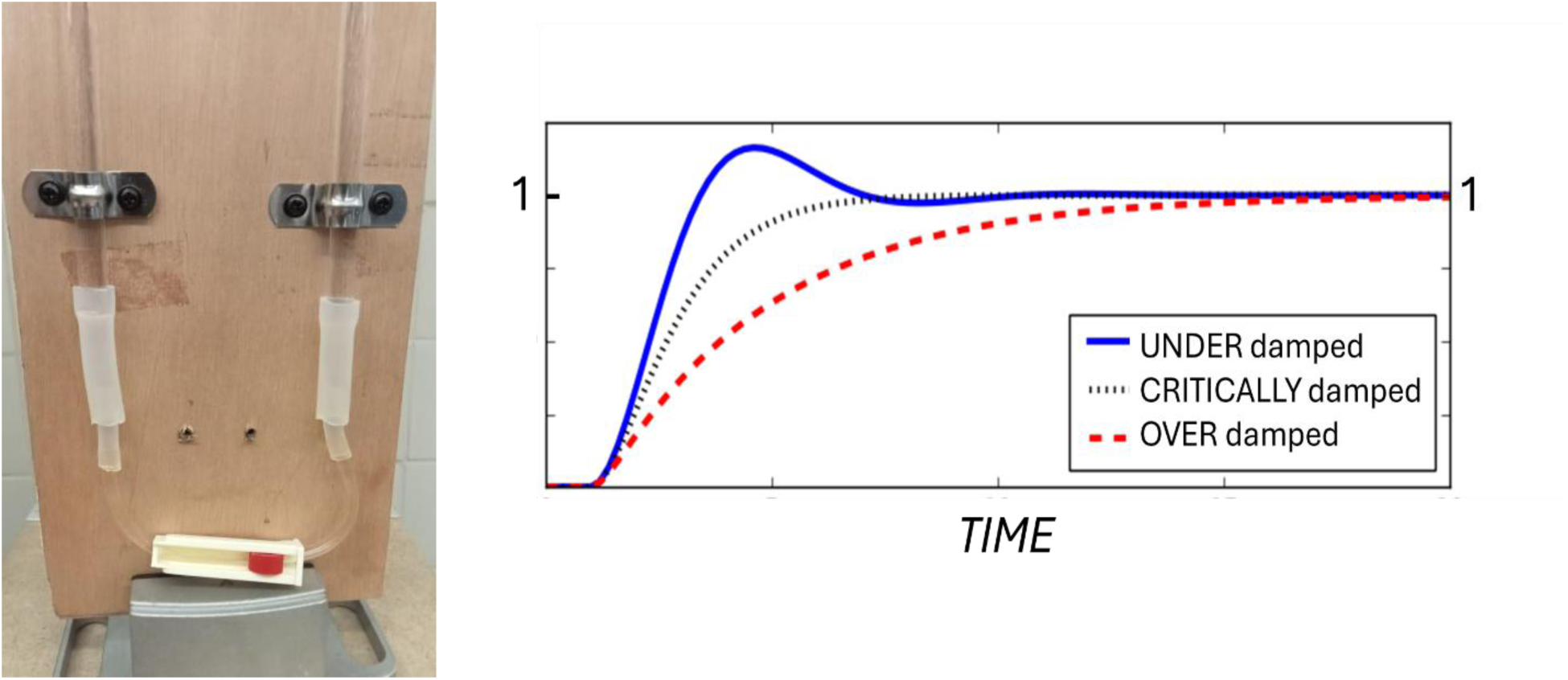
U-tube manometer. (A) Detail showing the roller clamp of a conventional intravenous infusion set placed in between the two branches. (B) Time course of the response of a second-order system when subjected to a step input of unit amplitude, for three cases: underdamped, critically damped and overdamped system.

Critical damping can be easily achieved through an experimental step-response procedure. First, a pressure difference is imposed between the two arms of the manometer, either by injecting air into one branch using a syringe or by laterally tilting the device, occluding one arm and subsequently restoring it to the vertical position. Upon sudden release of the pressure difference, the system response is observed. In an ideal system, the liquid column would return to equilibrium immediately. However, in the absence of additional resistance, the U-tube typically exhibits an underdamped oscillatory response (Figure 1.B). Progressive occlusion of the roller clamp increases flow resistance and reduces oscillations. When resistance is increased beyond the optimal value, the system becomes overdamped (Figure 1.B), resulting in a sluggish return to equilibrium. Thus, critical damping is achieved by gradually closing the roller clamp until oscillations are just eliminated (Figure 1.B). Although this adjustment can be performed by simple visual inspection (video tutorial in Supplementary materials), recording the step response using a smartphone camera and analyzing the video frame-by-frame will improve the accuracy of the measurement.

The performance of the critically damped U-tube manometer for monitoring peak inspiratory pressure was evaluated by comparison with a pressure transducer during mechanical ventilation of a patient model. To enhance measurement precision, the U-tube readings were video-recorded and observed frame-by-frame. Visual peak-pressure measurements obtained from the U-tube closely matched transducer readings, with deviations within 6%, for peak inspiratory pressures up to 30 cmH₂O and ventilation frequencies up to 30 breaths/min. At higher ventilation frequencies, however, the U-tube progressively underestimated peak pressure. For example, at 45 breaths/min –a high rate used only in selected pediatric cases– the underestimation reached approximately 30%. This reduction in accuracy at very high frequencies is consistent with the proximity of these ventilation rates to the intrinsic resonance frequency of the U-tube system. Therefore, the implemented manometer provides adequate accuracy for peak airway pressure monitoring in adult patients and in most pediatric ventilation settings.

### In vivo testing with a porcine model

*Animal preparation.* Four female Large White-Landrace pigs (48-52 kg) were obtained from an authorized supplier (Mir Ramadera, SL, Barcelona, Spain). The protocol was carried out with the required permission from the Animal Research Ethics Committee of the University of Barcelona (CEEA code: 694/25). Sedation was carried out with a mixture of xylazine 20% (2.2 mg/kg), ketamine 100 mg/mL (8 mg/kg), and midazolam 5 mg (0.6 mg/kg) administered intramuscularly, and anesthesia was induced with propofol (15 mg/kg). The animals were then orotracheally intubated with a 7.0 mm diameter tube with a suction port (Ruschalit, Teleflex, Ireland). Anesthesia was maintained by a continuous intravenous infusion of fentanyl (5µg/kg/h), midazolam (0.6 mg/kg/h) and propofol (8 mg/kg/h). Following intubation, the animals were initially ventilated using a SERVO-i mechanical ventilator (Maquet, Wayne, NJ, USA). Inspiratory gases were conditioned using a Conchatherm III humidifier, which was set to maintain the airway temperature near the Y-piece at 37°C. Airflow and airway pressure were recorded from a pneumotachograph and pressure transducers placed at the entrance of the endotracheal tube. Conventional mechanical ventilation was set as volume control, V_T_ = 8 ml/kg, FiO_2_ = 0.4, I:E ratio of 1:2, 10% inspiratory pause, PEEP= 4 cmH_2_O) and respiratory rate adjusted to maintain normocapnia. Arterial O_2_ saturation was non-invasively measured by pulse oximetry (Philips Efficia CM120 Patient Monitor) at the animal tail, and heart rate was monitored by ECG patches. Subsequently, under ultrasound guidance, the femoral artery was cannulated to obtain systemic arterial pressure with disposable transducers (TrueWave Pressure Transducer, Edwards Lifescience, Irvine, CA, USA) and to collect blood samples for arterial gasometry. Parameters were monitored by a Philips Efficia CM120 equipment. Finally, a size 12 Foley catheter was inserted into the bladder via vaginoscopy (3,4).

